# A plasmode simulation-based bias analysis for residual confounding by unmeasured variables leveraging information-rich subsets

**DOI:** 10.1101/2025.10.28.25338968

**Authors:** Rishi J Desai, Shirley V Wang, Haritha S. Pillai, Mufaddal Mahesri, Bowen Gu, Joyce Lii, Sarah Dutcher, Chanelle Jones, Fatma M. Shebl, Marie C. Bradley, Wei Hua, Hana Lee, Gerald J. Dal Pan, Sebastian Schneeweiss, Robert Ball

## Abstract

**Background:** Quantitative bias analyses often rely on unrealistic assumptions and do not fully reflect the complexities of healthcare data.

**Methods:** We describe a ‘plasmode’ simulation-based bias analysis for residual confounding from unmeasured variables by leveraging granular information from a subset of cohort members. We generated 500 simulated cohorts based on individual-level claims and linked electronic health record (EHR) data identifying new users of varenicline and bupropion from the Mass General Brigham site of the FDA Sentinel Real World Evidence Data Enterprise. Two adverse outcomes were simulated: 1) neuropsychiatric hospitalizations and 2) major adverse cardiovascular events (MACE), and measured confounding factors, identified from information available in claims including demographics, comorbid conditions, and comedications, were tailored to each outcome. Residual confounding was simulated using potential confounders measured in EHRs but unmeasured in claims including suicidal ideation for the neuropsychiatric outcomes and body mass index (BMI), blood pressure (BP), and smoking pack-years for the MACE outcome. These simulations retained the correlation between claims and EHR-based confounders observed in empirical data for realistic reflection of proxy adjustment of unmeasured confounders. Analyses were conducted in simulated data with and without adjustment for the EHR-based covariates to evaluate the extent of residual confounding in claims-only analyses.

**Results:** After 500 simulations, the median absolute standardized mean difference (ASMD) between treatment groups in the unadjusted sample was 0.16 for suicidal ideation; while <0.1 for BMI, BP, and smoking pack-years. For both outcomes, adjustment using claims-based variables provided relative bias close to 0, leading to the conclusion that EHR-measured confounders that were unmeasured in claims were unlikely to result in strong residual confounding within realistic simulations informed by empirical data.

**Conclusion:** The proposed approach provides a method for quantifying bias in non-randomized studies threatened by unavailability of potentially important confounding variables.

**Key points:**

- Residual confounding by unmeasured factors is a central threat in pharmacoepidemiology that is almost always acknowledged in published studies but seldom quantified.
- We describe a plasmode-simulation based approach to systematically design quantitative bias analyses that reflect the complexities of routinely collected healthcare data by leveraging detailed electronic health records from a subset.
- We provide open-source software code to enable other researchers to adopt this method in future studies and improve the reliability of their findings.

**Plain language summary:** This study introduces a new way for researchers to better understand and measure bias caused by missing health information in large insurance databases. Using detailed hospital records alongside insurance claims data, we created realistic computer simulations to test how much of the observed risk in safety studies could be explained away by missing important health factors, like depression or smoking habits, that aren’t always recorded in insurance data. The approach is flexible, uses real patient data, and helps researchers make stronger, more reliable conclusions about risks and benefits of treatments, even when some patient information is not available in all records.

## Introduction

Unmeasured confounders pose a central threat to the internal validity of non-randomized observational studies, but evaluation of such concerns quantitatively through bias analyses remains infrequent.^1^ Most studies that attempt to quantify the threat of confounding by unmeasured variables rely on simplistic methods that make many unrealistic assumptions.^1^ We recently proposed a flexible methodology based on individual-level data simulations that can allow researchers to relax many assumptions typically needed for bias analysis and characterize the bias arising from unmeasured confounders with a specified but modifiable structure.^2^

Here, we describe an extension that addresses a major limitation of the prior work, which was only designed to generate simplified scenarios with fully synthetic data involving a handful of measured confounders and did not simulate all correlations between measured confounders and unmeasured confounder(s). In real-world data, many more confounders may exist with correlations among them and with unmeasured confounders. Therefore, we now focus on implementation of more complex simulations in a ‘plasmode’ framework^3,4^ that uses actual individual-level data to preserve the naturally occurring correlations by resampling entire observations, keeping the covariate patterns intact. The treatment, the outcome, and their relationship are then simulated for the resampled observations based on observed covariates.^5^

The proposed approach is applicable in specific situations where investigators have access to measurements for a confounding variable of interest only in a subset of the overall population. An example that motivated the current work is in the context of the FDA Sentinel system, which is an active post marketing surveillance system for medical products in the US.^6^ The Sentinel distributed database (SDD) includes claims data from >200 million individuals across 14 data partners. Sentinel Real World Evidence Data Enterprise (RWE-DE) includes electronic health records (EHR) data linked with insurance claims data for >20 million individuals across six sites.^7^ Many studies investigating an exposure-outcome relationship in the SDD remain prone to residual confounding by factors unmeasured in claims data; however, in the smaller RWE-DE, some of these factors may be measured in a subset through EHRs, which could allow for supporting analyses to address uncertainties remaining with claims-only analyses. We use a specific case-example of the post-marketing safety of varenicline, a smoking cessation drug, to illustrate the methodology.

## Methods

### Case-example

We based the plasmode simulations on a realistic safety study that is reflective of typical investigations conducted in Sentinel. Varenicline had been hypothesized to increase the risk of adverse cardiovascular and neuropsychiatric outcomes. Numerous observational studies were conducted to generate evidence regarding safety of varenicline. For example, Pasternak et al.^8^ used national registry data from Denmark to compare risk of adverse neuropsychiatric events after treatment with varenicline versus bupropion. They noted that residual confounding by depressive symptoms, which may not be captured as structured diagnosis codes, could threaten the validity of their conclusion since bupropion may be selectively prescribed to those with underlying depression as it is also an antidepressant. Similarly, Graham et al.^9^ used Medicare claims data from the US to compare cardiovascular outcomes after treatment with varenicline versus bupropion and noted the potential threat of residual confounding by traditional cardiovascular risk factors such as smoking intensity and duration as well as body mass index, which are not recorded in claims data.

### Empirical study cohort

We used data from one of the RWE-DE sites,^7^ the Mass General Brigham (MGB), which contains EHR data from a large network of hospitals and outpatient clinics in the Boston metropolitan area linked to claims data from Medicaid (2000-2018) and Medicare (2007-2020) to identify a new-user cohort of patients dispensed varenicline or bupropion for smoking cessation. A 180-day baseline period prior to medication initiation was used to identify relevant confounders from claims data, which included demographics (age, race, sex), healthcare use factors (number of outpatient visits, emergency room visits, and hospitalizations), as well as comorbid conditions (focused on cardiometabolic conditions for the cardiovascular outcomes and psychiatric conditions for neuropsychiatric outcomes). We used claims-based outcome definitions that were previously reported by Pasternak et al.^8^ and Graham et al.^9^ for identifying adverse neuropsychiatric events and major adverse cardiac events (MACE), respectively.

### Information-rich subset

Among patients from the empirical study cohort identified in claims data, we derived detailed information on several potential confounding factors from EHRs in a subset where this information was available. For the neuropsychiatric outcome, severity of depression is a potential unmeasured confounder in claims data. Therefore, we extracted history of suicidal ideation as well as Patient Health Questionnaire (PHQ)-9 using natural language processing (NLP) of free-text notes. Notably, suicidal ideation is one of the most important features recognized in major depression severity scales.^10^ For the MACE outcome, potential confounders that are unmeasured in claims data include BMI, blood pressure and smoking intensity. We extracted BMI and blood pressure from structured EHR data and used NLP of free-text notes to extract pack-years to quantify smoking intensity and duration. Additional details regarding NLP methodology is provided in the **Appendix**. We described the characteristics of patients by whether they were included in the information-rich subset versus not, stratified by treatment.

#### Plasmode simulation workflow

We used the individual-level cohort data involving 21,964 new users of varenicline or bupropion for smoking cessation described above as the basis of the simulations. **Figure 1** summarizes the directed acyclic graphs informing data generation and **Figure 2** summarizes the key steps involved in the workflow. In the first step, we restrict the cohort to the subset of individuals who have recorded information in the EHR on the potential confounding variables of interest. For the neuropsychiatric outcome, this led to restriction to 18,696 individuals (85%) who had at least one free-text note in the baseline period for us to query for suicidal ideation and PHQ9 through NLP; for the MACE outcome, this led to restriction to 2,043 (9.3%) patients who had measurements on BMI, blood pressure, and smoking pack-years in the baseline period.

**Figure 1:**
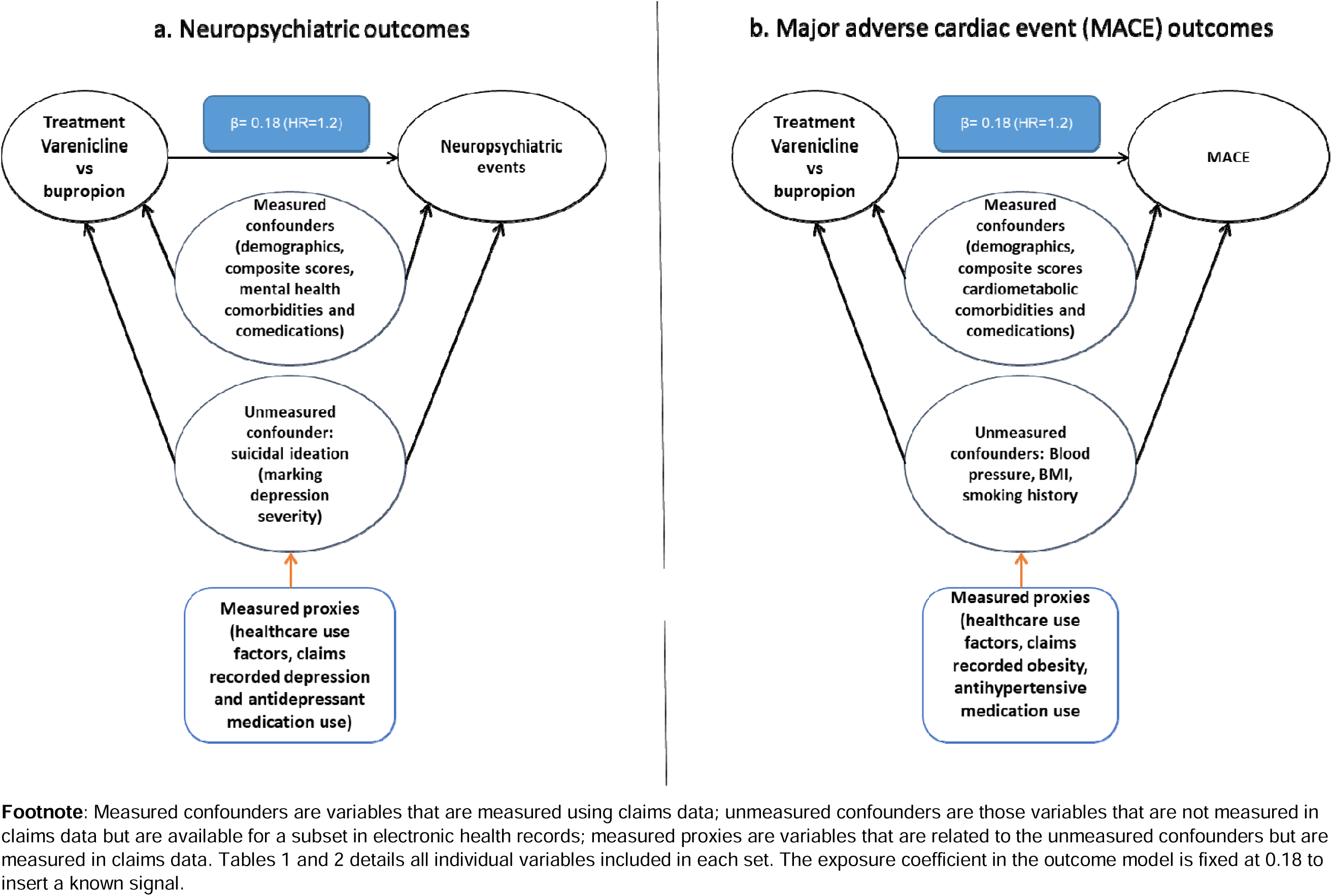
Directed acyclic graph for data generation Footnote: Measured confounders are variables that are measured using claims data; unmeasured confounders are those variables that are not measured in claims data but are available for a subset in electronic health records; measured proxies are variables that are related to the unmeasured confounders but are measured in claims data. Tables 1 and 2 details all individual variables included in each set. The exposure coefficient in the outcome model is fixed at 0.18 to insert a known signal.

**Figure 2:**
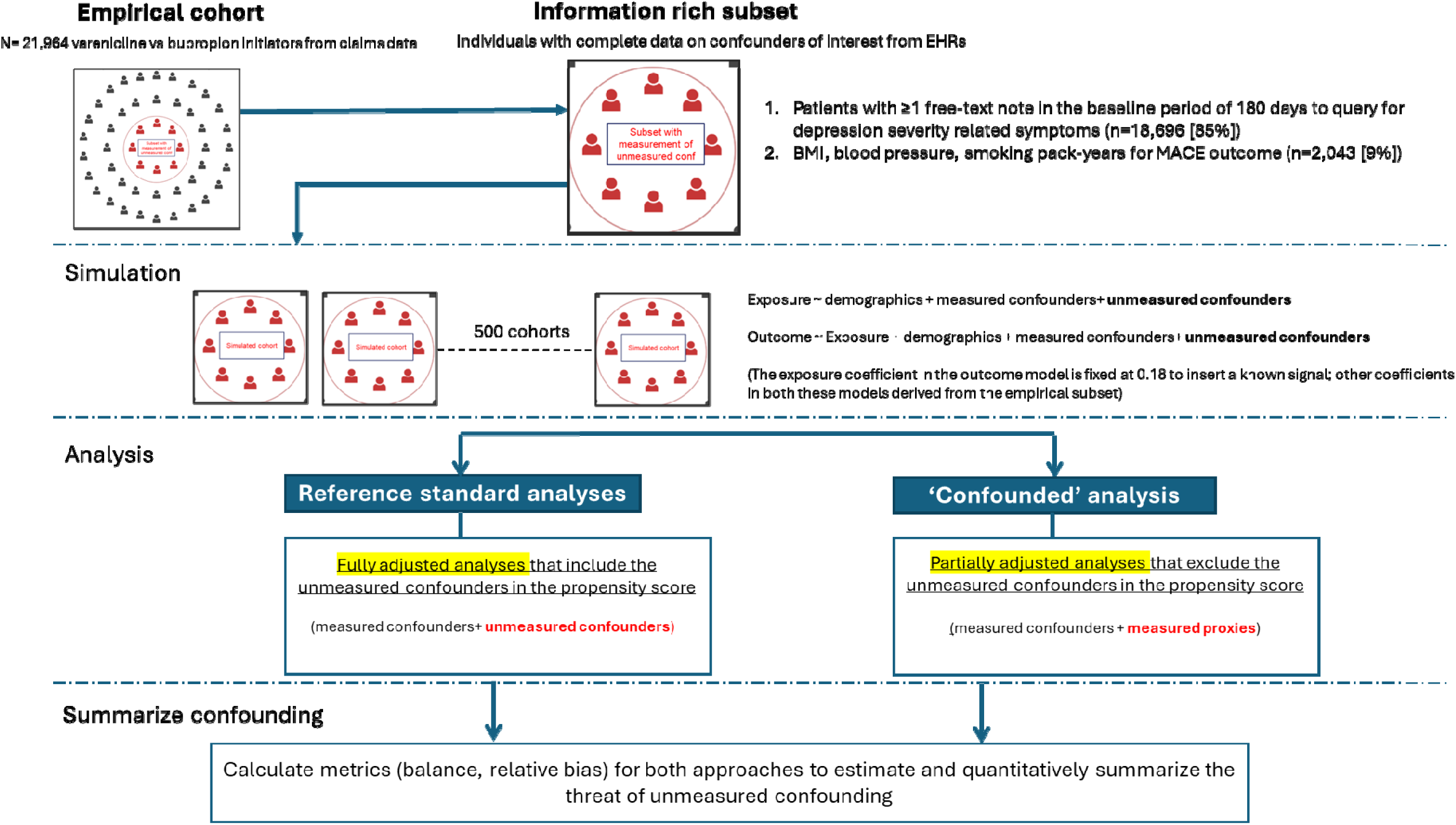
Plasmode simulation workflow.

**Table 1.**
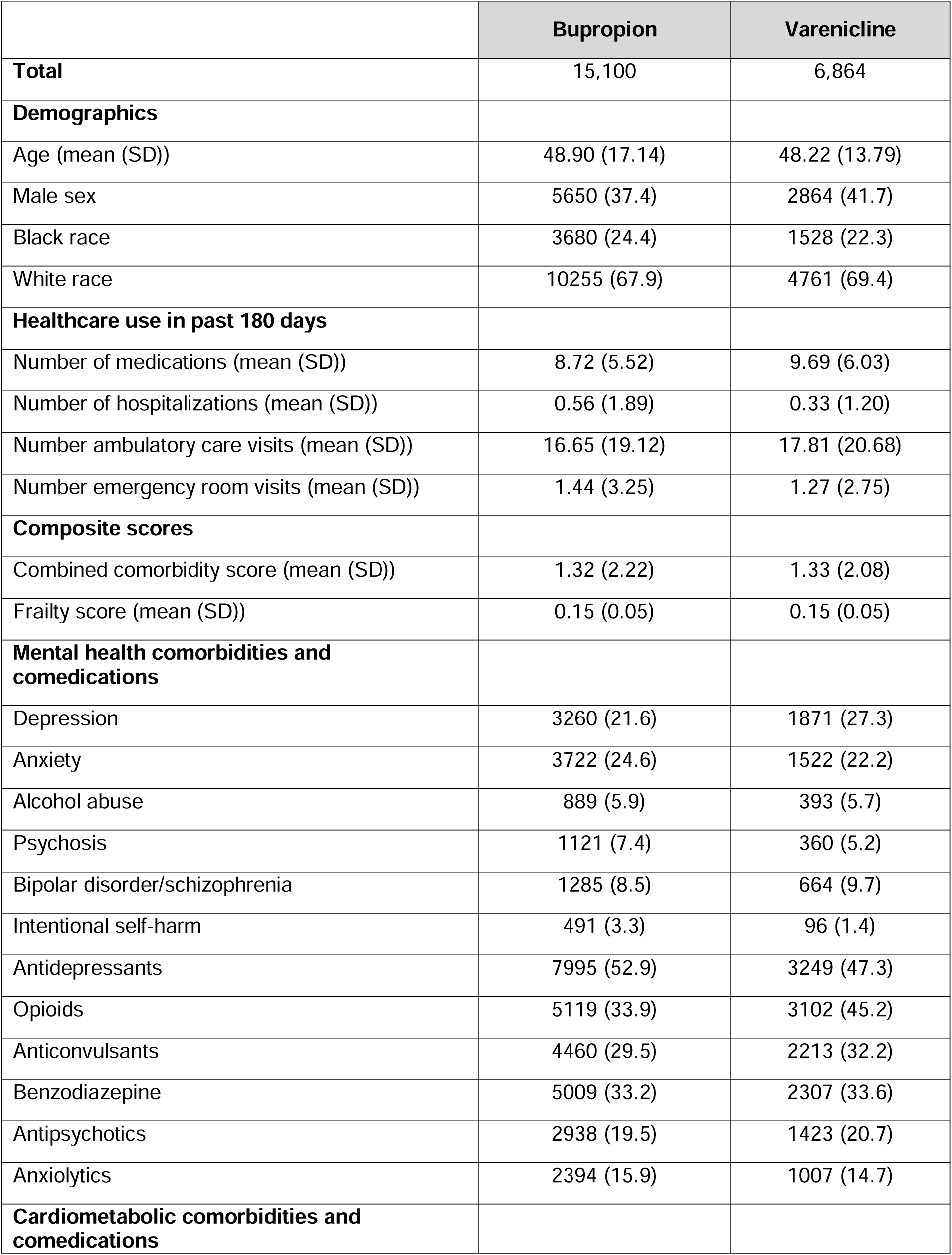
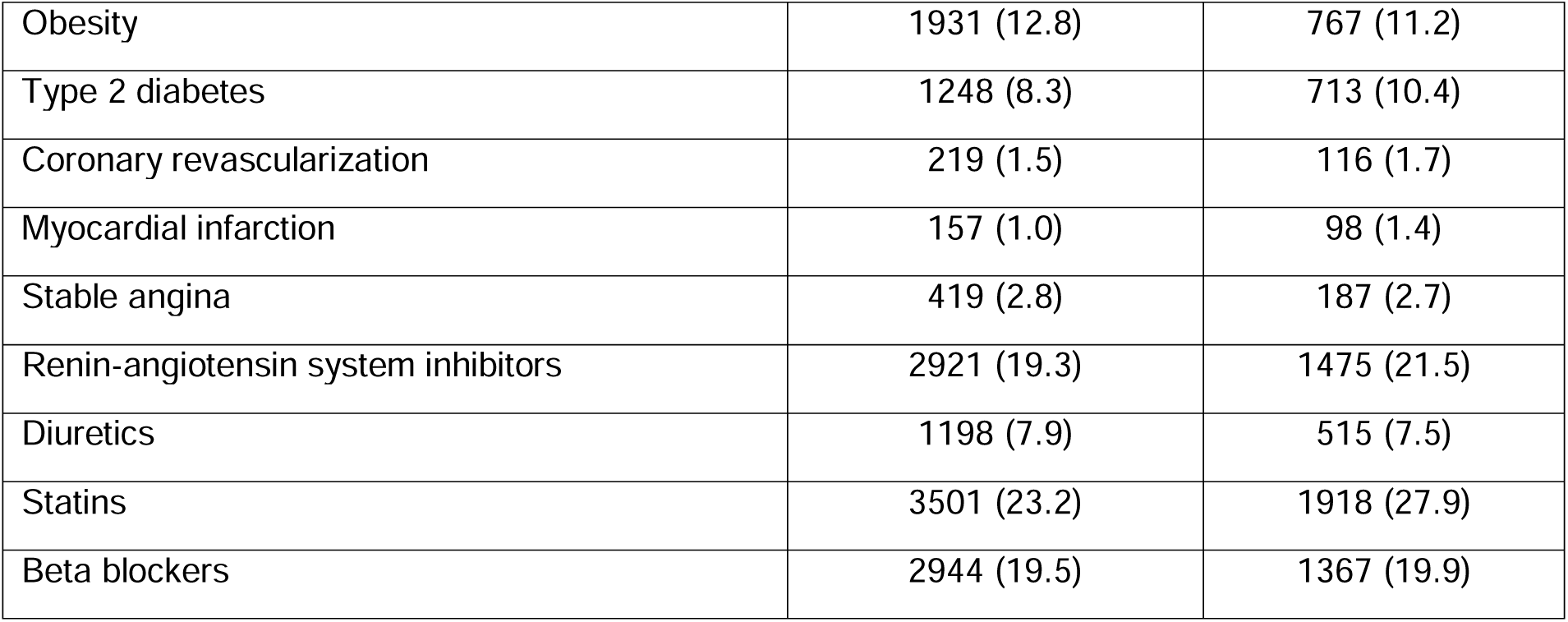
Patient characteristics measured in claims data for bupropion and varenicline initiators in the empirical study cohort.

|  | **Bupropion** | **Varenicline** |
| --- | --- | --- |
| **Total** | 15,100 | 6,864 |
| **Demographics** |  |  |
| Age (mean (SD)) | 48.90 (17.14) | 48.22 (13.79) |
| Male sex | 5650 (37.4) | 2864 (41.7) |
| Black race | 3680 (24.4) | 1528 (22.3) |
| White race | 10255 (67.9) | 4761 (69.4) |
| **Healthcare use in past 180 days** |  |  |
| Number of medications (mean (SD)) | 8.72 (5.52) | 9.69 (6.03) |
| Number of hospitalizations (mean (SD)) | 0.56 (1.89) | 0.33 (1.20) |
| Number ambulatory care visits (mean (SD)) | 16.65 (19.12) | 17.81 (20.68) |
| Number emergency room visits (mean (SD)) | 1.44 (3.25) | 1.27 (2.75) |
| **Composite scores** |  |  |
| Combined comorbidity score (mean (SD)) | 1.32 (2.22) | 1.33 (2.08) |
| Frailty score (mean (SD)) | 0.15 (0.05) | 0.15 (0.05) |
| **Mental health comorbidities and comedications** |  |  |
| Depression | 3260 (21.6) | 1871 (27.3) |
| Anxiety | 3722 (24.6) | 1522 (22.2) |
| Alcohol abuse | 889 (5.9) | 393 (5.7) |
| Psychosis | 1121 (7.4) | 360 (5.2) |
| Bipolar disorder/schizophrenia | 1285 (8.5) | 664 (9.7) |
| Intentional self-harm | 491 (3.3) | 96 (1.4) |
| Antidepressants | 7995 (52.9) | 3249 (47.3) |
| Opioids | 5119 (33.9) | 3102 (45.2) |
| Anticonvulsants | 4460 (29.5) | 2213 (32.2) |
| Benzodiazepine | 5009 (33.2) | 2307 (33.6) |
| Antipsychotics | 2938 (19.5) | 1423 (20.7) |
| Anxiolytics | 2394 (15.9) | 1007 (14.7) |
| **Cardiometabolic comorbidities and comedications** |  |  |
| Obesity | 1931 (12.8) | 767 (11.2) |
| Type 2 diabetes | 1248 (8.3) | 713 (10.4) |
| Coronary revascularization | 219 (1.5) | 116 (1.7) |
| Myocardial infarction | 157 (1.0) | 98 (1.4) |
| Stable angina | 419 (2.8) | 187 (2.7) |
| Renin-angiotensin system inhibitors | 2921 (19.3) | 1475 (21.5) |
| Diuretics | 1198 (7.9) | 515 (7.5) |
| Statins | 3501 (23.2) | 1918 (27.9) |
| Beta blockers | 2944 (19.5) | 1367 (19.9) |

**Table 2.** Information-rich subsets from electronic health records for bupropion and varenicline initiators in the empirical study cohort.

|  | **Bupropion** | **Varenicline** |
| --- | --- | --- |
| **Total** | 15,100 | 6,864 |
| **N with ≥1 free text note to query for relevant confounders for the neuropsychiatric outcome** | 12,747 (84.4%) | 5,849 (86.7%) |
| Suicidal ideation identified | 1,338 (10.5%) | 389 (6.6%) |
| PHQ-9 identified | 123 (1.0%) | 23 (0.4%) |
| PHQ-9 score (mean (SD)) | 13.0 (10.1) | 10.3 (6.7) |
| **N with information on relevant confounders for the MACE outcome** | 1,192 (7.9%) | 851 (12.4%) |
| Body mass index (mean (SD)) | 29.2 (7.5) | 28.9 (6.9) |
| Systolic blood pressure (mean (SD)) | 126.8 (18.0) | 126.9 (17.3) |
| Smoking history, pack-years (mean (SD)) | 22.4 (26.4) | 23.7 (25.8) |

The second step involves data simulation in the plasmode framework using the subset of patients with the ‘unmeasured’ confounders recorded. The plasmode simulation framework uses resampling with replacement to preserve realistic correlations between covariates when constructing simulated cohorts. In the resamples, we first simulated the binary treatment (varenicline or bupropion) using a logistic regression model including both claims-based and EHR-based patient characteristics. Coefficients used for treatment simulation were based on empirical associations between treatment initiation and these covariates estimated from the patient cohort described above. After simulating the treatment, we simulated the outcomes in the following steps. First, separate Cox proportional hazards models were specified to estimate multivariable associations for both claims-based and EHR-based patient characteristics with time to the outcome of interest and time to censoring. These models included an indicator for the treatment (varenicline or bupropion) to capture the relationships between covariates and the outcome (or censoring) independent of the treatment received. Next, we used the coefficient vectors from these survival models to predict event and censoring times for all patients using their covariate profiles. In this step, we replaced the treatment coefficient observed from empirical data with the desired treatment coefficient to simulate a known treatment effect. Finally, for each patient in each cohort, we simulated the follow-up time as the lesser of the predicted event time and the predicted censoring time. We defined the outcome as occurring if the predicted event time was less than predicted censoring time.

### Simulation studies

We separately explored the effect of unmeasured confounders for the MACE and neuropsychiatric events due to unique confounding structure for each outcome. Simulations for neuropsychiatric events used claims-based demographics, composite scores for comorbidity and frailty indices, as well as specific mental health related factors plus EHR-based suicidal ideation as confounding variables; while simulations for MACE used claims-based demographics, composite scores for comorbidity and frailty indices, as well as specific cardiometabolic factors listed in Table 1 plus EHR-based BMI, blood pressure, and smoking history measured by pack-years as confounding variables. Further, we specified several claims-based ‘proxy’ variables that could be related to the EHR-based variables. For neuropsychiatric events, the ‘proxy’ variables included claims-based depression diagnosis, antidepressant treatment, and healthcare utilization variables; for MACE, this included claims-recorded obesity diagnosis and healthcare utilization variables (number of medications, number of hospitalizations, emergency department visits, and outpatient visits). As shown in **Figure 1**, we did not use proxy variables in data generation, they were only used as ‘substitute’ variables for the EHR-based concepts in statistical analysis under the assumption that they are meaningfully correlated.

We kept the proportion of treatment and event rates for outcomes in each simulated cohort consistent with observed data. For both outcomes, 500 simulated cohorts were generated with sample sizes equal to the populations with complete measurement on EHR-based variables germane to each outcome of interest (18,696 for neuropsychiatric outcome; 2,043 for MACE). We simulated a true treatment effect of 1.2 on the hazard ratio scale (true coefficient = 0.18) for both outcomes.

### Statistical analysis

Following data generation, three levels of adjustments were used to estimate treatment effects, a) Level 0: no adjustment, b) Level 1: adjustment for claims-based confounders and claims-based proxies through propensity score weighting, c) Level 2: adjustment for both claims and EHR-based confounders through propensity score weighting. In all scenarios, we used logistic regression to model the PS and weighting reflecting the average treatment effect on the treated (“ATT” weighting, also referred to as weighting by the odds).^11^ Level 1 adjustment reflects commonly encountered situations where EHR-based information is not available, while Level 2 reflects ‘reference standard’ where residual confounding is completely mitigated in simulated data. Adjustment for claims-based variables in Level 1 can be expected to ameliorate confounding by EHR-based variables to varying degrees depending on the correlations between the claim-based proxies and EHR-based variables.

### Performance evaluation

To assess the degree of balance achieved in the EHR-based confounders with each level of adjustment, we plotted the distribution of average absolute standardized mean differences^12^ (ASMD) between treatment groups for these variables across 500 simulated cohorts for each outcome. Next, we calculated relative bias in treatment effect estimates with each level of adjustment on the hazard ratio scale as average of the difference between exponentiated estimated and exponentiated true coefficient divided by exponentiated true coefficient across 500 simulated cohorts in each scenario and presented as % bias.

#### R codes

Reusable R code and instructions for conducting tailored plasmode simulations based on user-specified parameters and dataset are publicly available.^13^ A file with instructions to use the code is provided in the **Appendix**.

## Results

### Empirical study cohort

**Table 1** summarizes patient characteristics among the two exposure groups in the empirical study cohort which formed the basis of our simulations. There were 15,100 bupropion initiators and 6,864 varenicline initiators. The average age (± standard deviation) was 49 ±17 years in the bupropion group and 48 ±14 years in the varenicline group. Among mental health comorbidities, the proportion with anxiety, psychosis, history of intentional self-harm, and use of antidepressants was higher among bupropion initiators; while among cardiometabolic comorbidities, the distribution was similar between the two groups.

### Information rich subsets

For the psychiatric outcome subset, we restricted to 18,696 individuals (85%) who had at least one free-text note in the baseline period for us to query for suicidal ideation through NLP and identified a history of suicidal ideation in 10.5% of the bupropion group and 6.6% of the varenicline group in this subset (**Table 2**). PHQ-9 was much less frequently recorded with only 146 total entries identified (<1%), so we could not use this variable further for simulations. For the MACE outcome subset, we restricted to a subset of 2,043 (9.3%) who had available information on all confounders of interest -BMI, blood pressure, pack-years-with average BMI slightly higher in bupropion group, while average smoking history was slightly longer for varenicline group.

**Appendix Tables 1 and 2** summarize characteristics of patients included in the subset and not included in the subsets, stratified by treatment, separately for the two outcomes. For the neuropsychiatric outcome subset, differences in patient characteristics were less prominent, between those included and not included in the subset; while for the MACE outcome subset, we observed that the patients included were older and had a greater burden of chronic comorbid conditions. We further noted that the general direction and magnitude of differences in patient characteristics between treatment groups were similar among patients included and not included in the subsets.

### Simulation results

Figure 3 shows results of simulations with respect to balance in the EHR-based variables with varying levels of adjustment. For the neuropsychiatric outcome, suicidal ideation was imbalanced between treatment groups with a median ASMD of 0.16 (interquartile range 0.14-0.18), which was reduced to 0.14 (interquartile range 0.12-0.16) with partial adjustment using claims-based proxies. For the MACE outcome, the imbalance in EHR-based variables was limited to begin with and became somewhat worse with partial adjustment but the medians remained <0.1 for all 3 variables.

**Figure 3:**
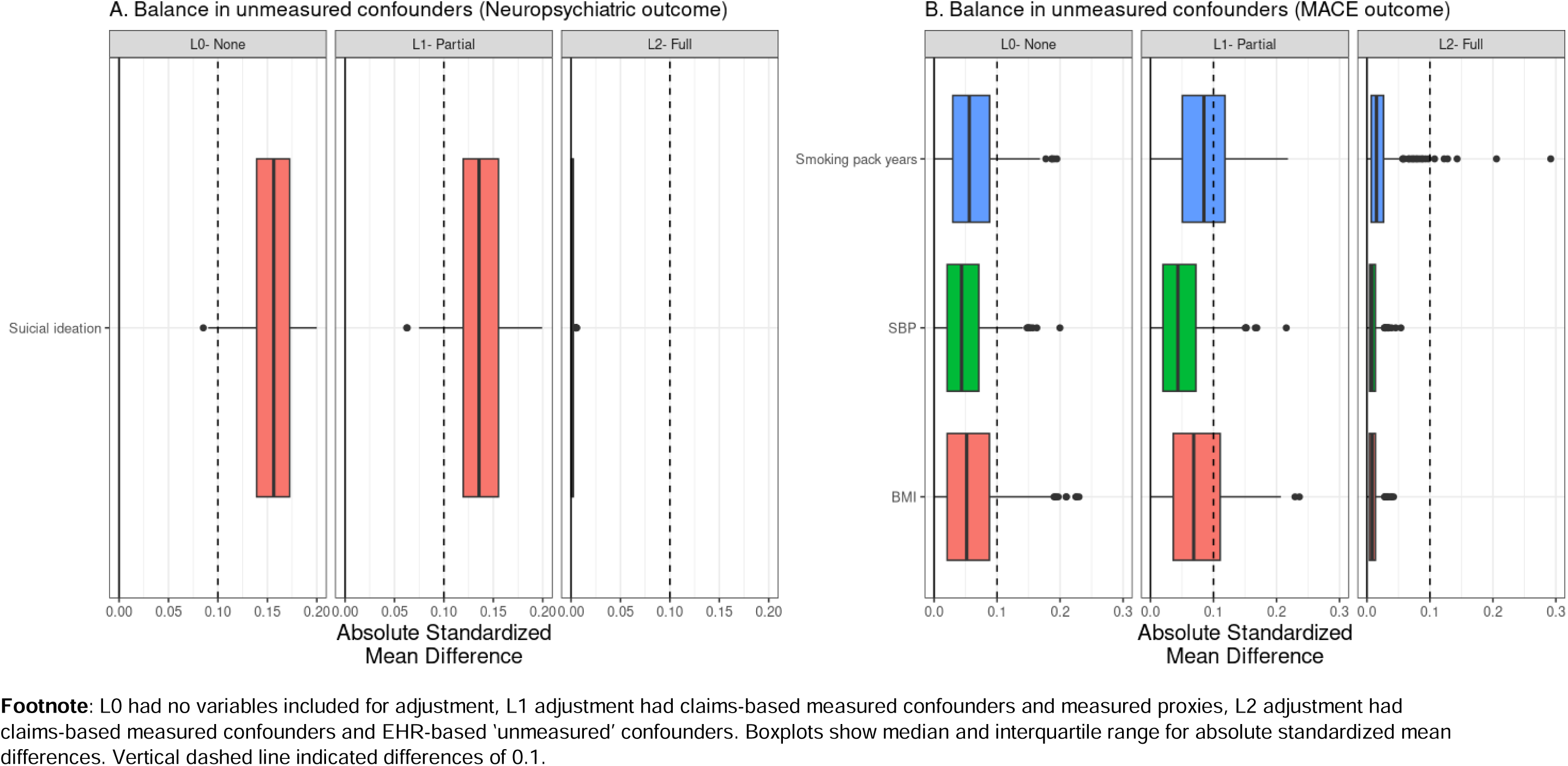
Balance in unmeasured confounders in 500 simulations with varying levels of confounding adjustment Footnote: L0 had no variables included for adjustment, L1 adjustment had claims-based measured confounders and measured proxies, L2 adjustment had claims-based measured confounders and EHR-based ‘unmeasured’ confounders. Boxplots show median and interquartile range for absolute standardized mean differences. Vertical dashed line indicated differences of 0.1.

Figure 4 shows the relative bias distribution across 500 simulations. For both outcomes, we noted some downward confounding with median relative bias of -3% and -13% for neuropsychiatric and MACE outcomes, respectively. We also observed that after partial adjustment with claims-based variables, the distributions moved quite close to 0 (median relative bias -0.4% and -1% for neuropsychiatric and MACE outcomes, respectively). Given these small residual relative bias values, we conclude that unmeasured factors that were proxy-adjusted but not directly measured in claims for both neuropsychiatric and MACE outcomes were unlikely to result in strong residual confounding within realistic simulated design and analytic scenarios informed by the empirical cohort.

**Figure 4:**
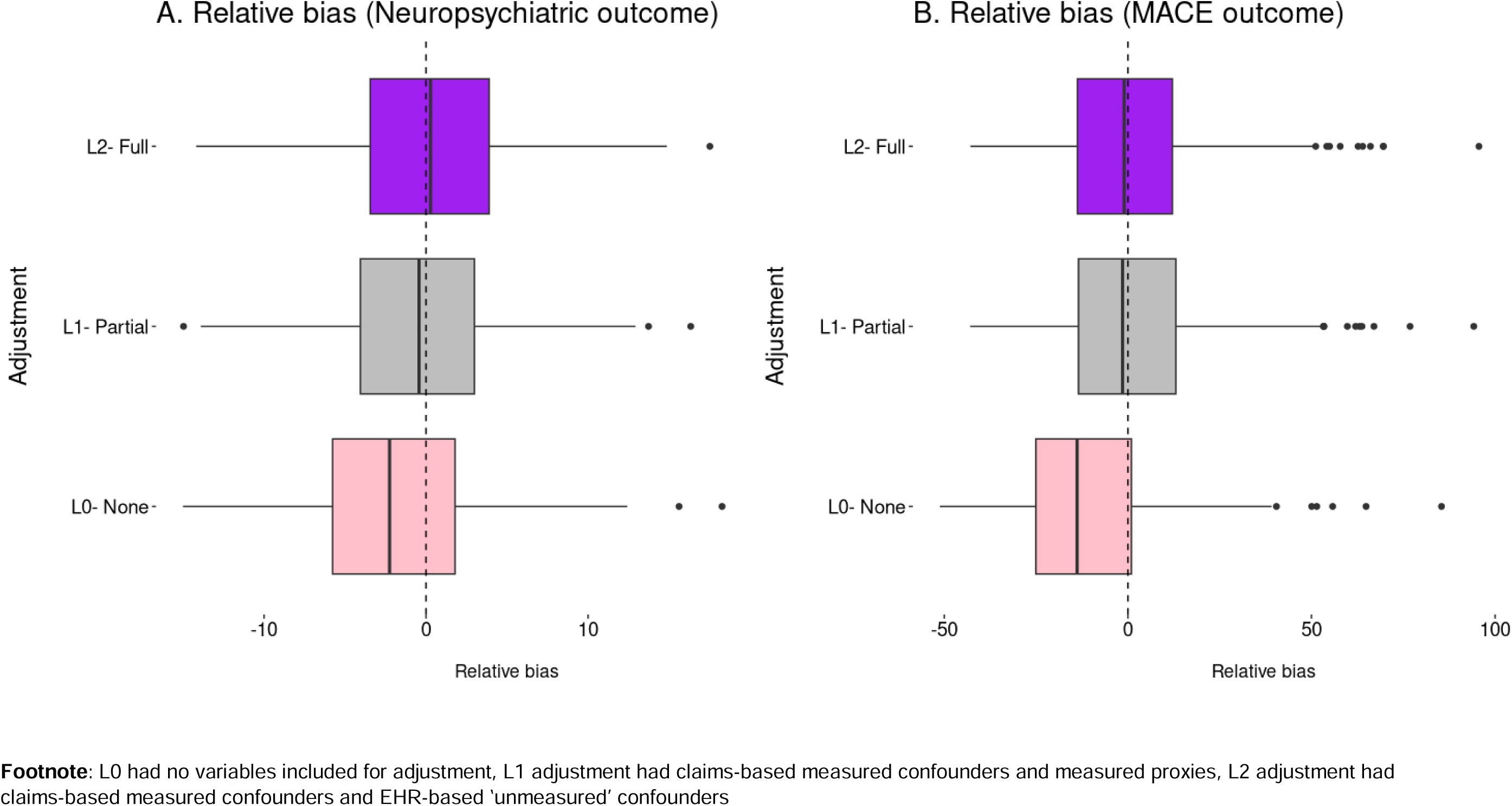
Relative bias across simulations with varying levels of confounding adjustment Footnote: L0 had no variables included for adjustment, L1 adjustment had claims-based measured confounders and measured proxies, L2 adjustment had claims-based measured confounders and EHR-based ‘unmeasured’ confounders

## Discussion

In the current study, we propose a methodology to leverage plasmode simulations and rich confounder information available from an enriched subset of the study cohort to provide a descriptive summary of the threat of residual confounding by unmeasured variables. We demonstrate the application of this methodology using a realistic case-example of the risk of neuropsychiatric and cardiovascular adverse events after treatment initiation with varenicline. The proposed methodology is flexible and easy to implement using software code that has been made available publicly.^13^

The concept of leveraging granular information from a subset of the cohort to address the threat of residual confounding has been explored in the literature. For instance, simple descriptive evaluations such as balance assessment between exposure groups in variables unmeasured in a larger cohort but available in a subset have been recommended previously.^14^ While balance assessment constitutes a good first step, this approach is limited because it remains unclear how the observed imbalance may impact effect estimates in specific circumstances. For instance, in our simulations, some imbalances remained in suicidal ideation for the neuropsychiatric outcome simulations after proxy adjustments; however, these imbalances had a limited impact on relative bias. In general, unmeasured variables that have substantial association with outcome risk and have limited related information recorded to stand in as proxy variables could pose greater threat to validity. By applying explicit simulation of treatment-outcome association under realistic scenarios using observed covariate patterns from real data, our approach is equipped to address this limitation of simple descriptive balance assessments.

The availability of granular information for a certain confounder only in a subset of the overall cohort can also be conceptualized as a missing data problem. Hence, statistical approaches to handle missing data may be considered as viable alternatives to adjust for variables that are only measured in a subset. For instance, multiple imputations, propensity score calibration, and inverse probability weighting approaches, including variations such as generalized raking that can calibrate the weights using information from auxiliary variables, have been shown to perform well in certain scenarios of partially observed confounder data, especially under missingness at random (MAR) assumption.^15–18^ However, when the proportion of missingness is large and data are missing not at random (MNAR), the performance is known to degrade with respect to bias in unpredictable ways. If investigators are uncertain about the missingness mechanism or are not willing to make the MAR assumption, our simulation-based approach provides an alternative. In real-world applications when data are MNAR, applying standard methods like imputations or calibration may provide biased results with a false reassurance of having dealt with the missingness. In contrast, the simulation-based approach is more descriptive and provides quantification of potential residual bias. In extreme circumstances when data are MNAR with strong value dependency, the simulation-based approach may be vulnerable to inadequate representation of bias because it starts out by restricting to complete cases. Notably, in these circumstances, standard methods (imputations, calibrations, or weighting) are also known to suffer from substantial bias.^17,18^ Therefore, we recommend conducting systematic evaluations of data missingness in the empirical sample on which simulations are based and reporting characteristics of patients included and not included in the subsets stratified by treatment to reassure readers. In our example, we observed that the subset with information on BMI, blood pressure, and smoking pack-years, had higher comorbidity burden and healthcare utilization compared to those who did not have this information. This is likely reflective of more intense monitoring and recording of information among these subjects because they are in more frequent contact with the healthcare system. However, we also observed that differences in patient characteristics between treatment groups followed largely similar patterns among subjects included and not included in the information-rich subsets. This observation suggests that the confounding structure may be preserved in the information rich subsets.

There are some salient points specific to the proposed approach that merit discussion. First, this approach is only applicable when investigators have access to information on the confounder(s) of interest in a subset of the cohort. When this is not the case, investigators may need to either use fully synthetic simulations we described previously^2^ or traditional methods for quantitative bias analysis that rely on aggregate level data.^19,20^ Second, while simulations based on empirical data provide a handy way to approximate reality, they may not always capture all the complexities observed in real data. Finally, when the subset with information on a critical confounder is small, as exemplified by the subset with information on PHQ9 scores in our example, it will be infeasible to proceed with this approach because sufficient empirical data are required to fit exposure and outcome models used for data generation in simulations.

In conclusion, we outline a plasmode simulation-based approach to quantify bias due to residual confounding using available data on unmeasured confounders from a subset. Information generated from simulations can help with interpretation of results of observational studies potentially threatened by residual confounding.

## Supporting information

plasmode_simba_demo

## Data Availability

All data produced in the present study are available upon reasonable request to the authors

file:///C:/Users/hp814/AppData/Local/Microsoft/Windows/INetCache/Content.Outlook/EIRXYKJU/Appendix%20-%20plasmode_simba%20demo.html

