## supplemental material for "A plasmode simulation-based bias analysis for residual confounding by unmeasured variables leveraging information-rich subsets"

**Appendix methods**

*Extraction of additional confounders from free-text notes using natural language processing*

The smoking pack years, suicidal ideation, and PHQ-9 scores were extracted by applying regular expressions to the EHR notes collected during the 180-day baseline period. Specifically, the smoking pack years were matched in two different ways: matching the explicit mention of pack years (Type I) or matching the mention of the smoke years together with the packs per day (Type II). The suicidal ideation was matched with a set of regular expressions described in the table below, and the notes were considered to indicate suicidal ideation if any of these regular expressions are matched. The PHQ-9 scores were matched with a set of regular expressions described in the table below, and the PHQ-9 scores were directly extracted from the matching results.

| **Variable** | **Query terms** |
| --- | --- |
| **Suicidal ideation** | suicid(al\|e) attempt |
|  | suicid(al\|e) ideation and attempt |
|  | (attempted\|committed) suicide |
|  | (try\|tried\|tries\|trying\|attempted\|attempts\|attempting) (of\|to) (take\|end) (ing) (my\|his\|her\|their) (own) life |
|  | (try\|tried\|tries\|trying\|attempted\|attempts\|attempting) (of\|to) (kill\|shot\|shoot\|hang\|poison\|asphyxiate\|asphyxiat\|mutilate\|mutilat\|harm\|overdose\|overdos\|cut\| cutt\|gas\|gass\|slash) (ing)* (myself\|himself\|herself\|themself) |
|  | (try\|tried\|tries\|trying\|attempted\|attempts\|attempting) (of\|to) (slit\|slitt\|cut\|cutt\|slash) (ing) (my\|his\|her\|their\|the) (wrist\|arm\|throat) |
|  | (try\|tried\|tries\|trying\|attempted\|attempts\|attempting) (of\|to) (jump\|jumping) off (a\|the\|interstate\|my\|his\|her\|their) (bridge\|building\|balcony\|window\|roof) |
|  | (try\|tried\|tries\|trying\|attempted\|attempts\|attempting) (of\|to) (jump\|jumping) out of (a\|the) moving (vehicle\|car) |
|  | (try\|tried\|tries\|trying\|attempted\|attempts\|attempting) (of\|to) (jump\|jumping) from a moving (vehicle\|car) |
|  | (try\|tried\|tries\|trying\|attempted\|attempts\|attempting) (of\|to) (jump\|jumping) out of (his\|her\|the\|a) (\d+) (nd\|rd\|th) (floor\|story\|balcony\|window) |
|  | (try\|tried\|tries\|trying\|attempted\|attempts\|attempting) (of\|to) (jump\|jumping) in front of a (car\|truck\|train\|vehicle) |
|  | (try\|tried\|tries\|trying\|attempted\|attempts\|attempting) (of\|to) (jump\|jumping) into interstate |
|  | (try\|tried\|tries\|trying\|attempted\|attempts\|attempting) (of\|to) (jump\|jumping) out of (a\|the\|his\|her) (window\|balcony) |
| **PHQ9 score** | 'PHQ[-\s]?9[:\s]*(score)?[:\s]*(\d{1,2})' |

After getting the extraction results of each EHR note, the note-level results were aggregated to the patient-level results based on the following rules:

Pack Years: If only one unique value is extracted, then we used this value. If multiple unique values were extracted, then we used the largest value extracted using Type I matching, or the largest value extracted using Type II matching if no Type I matching results were available. If no values were extracted, then we considered this information to be not available.

Suicidal Ideation: Suicidal ideation was identified as “yes” if any note has the suicidal ideation regular expressions matched, otherwise no.

PHQ-9: If only one unique value was extracted, then use this value. If multiple unique values were extracted, then we used the largest extracted value. If no values were extracted, then we considered this information to be not available.

**Appendix Table 1: Characteristics of patients included in the subset and not included in the subset for the MACE outcome simulations, stratified by treatment**

|  | **Subset with additional confounder information** | | | **Subset without additional confounder information** | | |
| --- | --- | --- | --- | --- | --- | --- |
|  | Varenicline | Bupropion | Total | Varenicline | Bupropion | Total |
| n | 851 | 1192 | 2043 | 6013 | 13908 | 19921 |
| **Demographics** |  |  |  |  |  |  |
| Age (mean (SD)) | 55.06 (12.96) | 57.54 (15.52) | 56.51 (14.55) | 47.25 (13.64) | 48.16 (17.07) | 47.88 (16.12) |
| Male sex | 47% | 38% | 42% | 41% | 37% | 38% |
| White race | 73% | 72% | 73% | 69% | 67% | 68% |
| **Healthcare use factors** |  |  |  |  |  |  |
| Number of medications (mean (SD)) | 10.79 (5.98) | 10.23 (5.65) | 10.47 (5.79) | 9.54 (6.02) | 8.59 (5.49) | 8.88 (5.67) |
| Number of hospitalizations (mean (SD)) | 0.38 (0.93) | 0.61 (1.43) | 0.51 (1.25) | 0.33 (1.24) | 0.56 (1.92) | 0.49 (1.75) |
| Number ambulatory care visits (mean (SD)) | 16.53 (18.21) | 18.49 (19.00) | 17.67 (18.70) | 17.99 (21.00) | 16.49 (19.12) | 16.94 (19.72) |
| Number emergency room visits (mean (SD)) | 1.05 (2.14) | 1.36 (2.78) | 1.23 (2.54) | 1.30 (2.82) | 1.45 (3.28) | 1.40 (3.15) |
| **Composite scores** |  |  |  |  |  |  |
| Combined comorbidity score (mean (SD)) | 2.23 (2.61) | 2.55 (3.12) | 2.42 (2.92) | 1.20 (1.96) | 1.21 (2.09) | 1.21 (2.05) |
| Frailty score (mean (SD)) | 0.16 (0.05) | 0.17 (0.06) | 0.17 (0.05) | 0.14 (0.05) | 0.15 (0.05) | 0.15 (0.05) |
| **Cardiometabolic comorbid conditions and comedications** |  |  |  |  |  |  |
| Obesity | 19% | 23% | 21% | 10% | 12% | 11% |
| Type 2 diabetes mellitus | 20% | 18% | 19% | 9% | 7% | 8% |
| Coronary revascularization | 3% | 5% | 4% | 1% | 1% | 1% |
| Myocardial infarction | 2% | 3% | 3% | 1% | 1% | 1% |
| Stable angina | 6% | 8% | 7% | 2% | 2% | 2% |
| ACE/ARB use | 30% | 31% | 31% | 20% | 18% | 19% |
| Diuretics | 13% | 14% | 13% | 7% | 7% | 7% |
| Statins | 44% | 42% | 43% | 26% | 22% | 23% |
| Beta blockers | 27% | 32% | 30% | 19% | 18% | 19% |

**Appendix Table 2: Characteristics of patients included in the subset and not included in the subset for the neuropsychiatric adverse outcome simulations, stratified by treatment**

|  | **Subset with additional confounder information** | | | **Subset without additional confounder information** | | |
| --- | --- | --- | --- | --- | --- | --- |
|  | Varenicline | Bupropion | Total | Varenicline | Bupropion | Total |
| n | 5949 | 12747 | 18696 | 914 | 2316 | 3230 |
| **Demographics** | 47.87 (13.75) | 48.74 (17.06) | 48.46 (16.08) | 50.51 (13.88) | 49.93 (17.60) | 50.09 (16.63) |
| Age (mean (SD)) | 42% | 37% | 39% | 38% | 36% | 37% |
| Male sex | 68% | 67% | 67% | 76% | 74% | 75% |
| White race |  |  |  |  |  |  |
| **Healthcare use factors** | 9.63 (5.99) | 8.71 (5.54) | 9.00 (5.70) | 10.08 (6.22) | 8.77 (5.45) | 9.14 (5.71) |
| Number of medications (mean (SD)) | 0.34 (1.20) | 0.58 (1.89) | 0.50 (1.70) | 0.26 (1.21) | 0.39 (1.37) | 0.35 (1.33) |
| Number of hospitalizations (mean (SD)) | 17.84 (20.69) | 16.82 (19.42) | 17.14 (19.84) | 17.56 (20.50) | 15.37 (16.69) | 15.99 (17.87) |
| Number ambulatory care visits (mean (SD)) | 1.30 (2.78) | 1.47 (3.30) | 1.42 (3.15) | 1.06 (2.51) | 1.13 (2.32) | 1.11 (2.37) |
| Number emergency room visits (mean (SD)) |  |  |  |  |  |  |
| **Composite scores** | 1.32 (2.09) | 1.33 (2.23) | 1.33 (2.18) | 1.39 (2.00) | 1.23 (2.14) | 1.27 (2.11) |
| Combined comorbidity score (mean (SD)) | 0.15 (0.05) | 0.15 (0.05) | 0.15 (0.05) | 0.15 (0.05) | 0.14 (0.05) | 0.14 (0.05) |
| Frailty score (mean (SD)) | 5949 | 12747 | 18696 | 914 | 2316 | 3230 |
| **Mental health comorbidities and comedications** |  |  |  |  |  |  |
| Depression | 27% | 22% | 23% | 29% | 20% | 22% |
| Anxiety | 21% | 24% | 23% | 29% | 26% | 27% |
| Alcohol abuse | 5% | 6% | 6% | 7% | 6% | 6% |
| Psychosis | 5% | 7% | 7% | 6% | 6% | 6% |
| Bipolar disorder/schizophrenia | 9% | 8% | 9% | 12% | 9% | 10% |
| Intentional self-harm | 1% | 3% | 3% | 1% | 2% | 2% |
| Antidepressants | 47% | 52% | 51% | 49% | 56% | 54% |
| Opioids | 45% | 34% | 38% | 45% | 32% | 36% |
| Anticonvulsants | 31% | 29% | 30% | 37% | 31% | 33% |
| Benzodiazepine | 33% | 33% | 33% | 35% | 34% | 34% |
| Antipsychotics | 20% | 19% | 19% | 24% | 21% | 22% |
| Anxiolytics | 14% | 16% | 15% | 16% | 16% | 16% |
