## Supplementary material for "A plasmode simulation-based bias analysis for residual confounding by unmeasured variables leveraging information-rich subsets": plasmode_simba_demo: Appendix - plasmode_simba demo.html


### plasmode\_simba demo

###### Rishi Desai

#### plasmode\_simba

Unmeasured confounding is often raised as a source of potential bias
when evaluating non-randomized study protocols, but evaluating such
concerns quantitatively remains challenging and dependent on simplistic
methods that need make many unrealistic assumptions. We recently
proposed a flexible methodology based on individual level simulations
that can allow researchers to characterize the bias arising from
unmeasured confounding with a specified but modifiable structure during
the study design. Our package “sim.BA” allows user to conduct a
simulation-based quantitative bias analysis using covariate structures
generated with individual-level data to characterize the bias arising
from unmeasured confounding.

Functions included in this extension allow users to implement complex
simulations that represent a realistic description of the relationships
among multiple confounders and outcomes in a ‘plasmode’ framework that
builds on actual patient-level data as the basis of simulations to
preserve the naturally occurring correlation patterns among study
variables.

#### Basic set up

As plasmode\_simba function uses actual patient data as seed to
conduct simulations, the first requirement is availability of patient
level analytic file with information on exposure, outcome, survival
time, measured and unmeasured confounders. In this demo, we have the
file ‘cohort’ as our patient level analytic file (“input\_data”) that
forms the basis for simulation.

```
#Include all the functions and files needed for simulation

source("~/simBA_plasmode/functions/Analysis_function_final.R")

source("~/simBA_plasmode/functions/plasmode_simba.R")

source("~/simBA_plasmode/functions/plot.plasmode_simba.R")

cohort <- read.csv("~/simBA_plasmode/MACE_final_sample.csv") #this is the 'complete-case' input data with information on EHR based variables
```

Next, you will need to create a simple input table tagging which
variables in your seed data correspond to which role (possible roles
include: exposure, survtime, outcome, measured\_confounder,
unmeasured\_confounder, proxy). For the ‘cohort’ dataset, following is
the table that we will use as our ‘input file’

```
input_file <- read.csv("~/simBA_plasmode/input_varenicline_MACE.csv")

print(input_file)
#>                      Variable                  Role
#> 1                 varenicline              exposure
#> 2                followuptime              survtime
#> 3                     outcome               outcome
#> 4                         Age   measured_confounder
#> 5                         sex   measured_confounder
#> 6                        Race   measured_confounder
#> 7                  NumGeneric                 proxy
#> 8                       NumIP                 proxy
#> 9                       NumAV                 proxy
#> 10                      NumED                 proxy
#> 11                        CCI   measured_confounder
#> 12                    FRAILTY   measured_confounder
#> 13                    Obesity                 proxy
#> 14                       T2DM   measured_confounder
#> 15 Coronary_revascularization   measured_confounder
#> 16       Myocardial_infaction   measured_confounder
#> 17              Stable_angina   measured_confounder
#> 18                    ACE_ARB                 proxy
#> 19                  Diuretics                 proxy
#> 20                    Statins   measured_confounder
#> 21              Beta_blockers                 proxy
#> 22                  Vital_BMI unmeasured_confounder
#> 23             Vital_Systolic unmeasured_confounder
#> 24       Pack_Year_Aggregated unmeasured_confounder
```

Following is the directed acyclic graph used for data generation in
this example.

Simulations used claims-based demographics, composite scores for
comorbidity and frailty indices, as well as specific cardiometabolic
factors plus EHR-based BMI, blood pressure, and smoking history measured
by pack-years as confounding variables contributing to both exposure and
outcome models. Additionally, exposure variable is included in the
outcome model. Further, we specified several claims-based ‘proxy’
variables that are presumably related to the EHR-based variables.This
included claims-recorded obesity diagnosis and healthcare utilization
variables (number of medications, number of hospitalizations, emergency
department visits, and outpatient visits). Notably, proxy variables are
not entered in either exposure or outcome models for data generation,
they are only meant to be used as ‘substitute’ variables for the
EHR-based concepts in statistical analysis under the assumption that
they are meaningfully correlated.

#### Running simulations

```
results <- plasmode_simba (input_data= cohort, id="PatID", treatment_effect= 1.2, num_obs= 2043, iterations=500, 
                           input_file=input_file,
                           adj="weighting", adj_args= list(method = "glm", 
                                                           estimand="ATT"))
```

plasmode\_simba is the flagship function which calls in other
functions to do the following key steps based on the information
provided in the input file

1. Uses the patient level input data with information on all study
   variables (exposure, outcome, survival time, measured, and unmeasured
   confounders) to generate plasmode simulated datasets. In practice, this
   patient level input data may be a subset (e.g subjects with EHR linked
   data) from an overall cohort (e.g arising from insurance claims data)
   where more granular information on confounders is recorded, which remain
   unmeasured in the overall cohort. In the plasmode simulations, exposure
   and time-to-event outcomes are generated based on logistic and cox
   models, respectively. Variables listed as confounders (measured or
   unmeasured) in the input file are included in these models with
   coefficients derived from the patient level input data.
2. Once the data are simulated, the function moves on to the analysis
   step. A total of 3 Cox proportional hazard regression models are
   implemented: 1) a crude model that does not include any confounders, 2)
   partially adjusted analyses (L1) that only includes measured confounders
   and proxies, 3) fully adjusted analyses (L2) that include both measured
   and unmeasured confounders. For adjustment, users can implement
   propensity score based approach of their choice (matching, weighting,
   subclassification etc). The intuition here is that the partially
   adjusted analyses (L1) can be considered ‘confounded’ because it does
   not include known unmeasured confounders, and the magnitude of the
   unmeasured confounding can be gaged by comparing (L1) results to (L2)
   results, which serve as the reference standard.
3. The output from the plasmode\_simba function is a list including 2
   tables: 1) a table that contains balance measures (absolute standardized
   mean difference) for the variables listed as unmeasured confounders with
   each level of adjustment, 2) results for each individual simulation run
   including hazard ratios, standard errors, and percent bias.

The following table provides guidance on how to design your
simulations using various options available in the plasmode\_simba ( )
function.

| Option | Specification notes |
| --- | --- |
| input\_data | REQUIRED: the patient level analytic file with information on all study variables (at a minimum: exposure, outcome, survival time, measured confounder(s), unmeasured confounder(s)) |
| input\_file | REQUIRED: input table tagging which variables in your seed data correspond to which role (possible roles include: exposure, survtime, outcome, measured\_confounder, unmeasured\_confounder, proxy (optional)) |
| id | REQUIRED: variable indicating unique patient identifier in the input\_data |
| treatment\_effect | REQUIRED: desired treatment effect to be simulated |
| num\_obs | REQUIRED: total number of observations per simulated dataset |
| iterations | REQUIRED: total number of simulation runs |
| adj | REQUIRED: Adjustment method, options include “matching” or “weighting” on the propensity score. |
| adj\_args | REQUIRED: A list of arguments passed to `MatchIt::matchit()` or `WeightIt::weightit()` depending on the argument to `adj`. Take care to specify these arguments to ensure the adjustment method is as desired. |

#### Reviewing results

```
plot.plasmode_simba(results, "balance")
```

```
plot.plasmode_simba(results, "bias")
```

```
plot.plasmode_simba(results, "hr")
```

The results can be visualized as boxplots showing distribution
balance as well as relative bias with each level of adjustment. If
distribution of relative bias is similar for L1 and L2 adjustments, it
provides some indication that the threat of unmeasured confounding is
likely not very severe. In case of notable differences in distribution
between L1 and L2 adjustments, one would conclude that the threat of
unmeasured confounding is likely severe.

```
pb_table <- results[["HR_summary"]] %>% group_by(adjustment) %>% 
  summarise(across(c(pbias),median),
            .groups = 'drop') %>%
  as.data.frame()

pb_table
#>    adjustment     pbias
#> 1    L0- None -9.737654
#> 2 L1- Partial  1.447534
#> 3    L2- Full  1.192920


HR_table <- results[["HR_summary"]] %>% group_by(adjustment) %>% 
  summarise(across(c(expHR),median),
            .groups = 'drop') %>%
  as.data.frame()

HR_table
#>    adjustment    expHR
#> 1    L0- None 1.104694
#> 2 L1- Partial 1.225093
#> 3    L2- Full 1.225108

SE_table <- results[["HR_summary"]] %>% group_by(adjustment) %>% 
  summarise(across(c(SE),mean),
            .groups = 'drop') %>%
  as.data.frame()

SE_table
#>    adjustment        SE
#> 1    L0- None 0.1069314
#> 2 L1- Partial 0.1161941
#> 3    L2- Full 0.1161983
```

One could also summarize means or medians for hazard ratios, standard
errors, or relative bias across simulation runs to compare these values
across different adjustment strategies.
